# Further analysis of the impact of distancing upon the COVID-19 pandemic

**DOI:** 10.1101/2020.04.14.20048025

**Authors:** Daniel J. Bernstein

## Abstract

This paper questions various claims from the paper “Social distancing strategies for curbing the COVID-19 epidemic” by Kissler, Tedijanto, Lipsitch, and Grad: most importantly, the claim that China’s “intense” distancing measures achieved only a 60% reduction in *R*_0_.

## 1 Introduction

The 22 March 2020 paper “Social distancing strategies for curbing the COVID-19 epidemic” [**5**] reports calculations in a model where distancing reduces *R*_0_ by at most 60%, and claims that 60% is “on par with the reduction in *R*_0_ achieved in China through intense social distancing measures (3)”.

Reference “(3)” is a 9 March 2020 paper [**1**] that does not say what [**5**] claims it says. What [**1**] actually says about the effect of China’s distancing upon *R*_0_ is the following:

> With an early epidemic value of *R*_0_ of 2.5, social distancing would have to reduce transmission by about 60% or less, if the intrinsic transmission potential declines in the warm summer months in the northern hemisphere. This reduction is a big ask, but it did happen in China.

In other words, if *R*_0_ in China was originally 2.5 without distancing, and if China’s interventions had less than a 60% effect, then the new *R*_0_ would have been larger than 1, so the epidemic would have continued to spread exponentially in China—but the news reports say that the epidemic was stopped in China, so *R*_0_ there must have been reduced by *at least* 60%.

How does [**5**] conclude that “intense” distancing reduces *R*_0_ by *at most* 60%? There do not seem to be any further comments on this topic in [**5**]; also, [**1**] does not seem to present any justification for its claim that reducing *R*_0_ by 60% is a “big ask”. Is it plausible that an “intense” lockdown—presumably a drastic reduction in the amount of contact between typical people—would reduce transmission by only 60%? The actual level of reduction directly affects the main quantitative conclusions of [**5**].

The main objective of this paper is to see what the distancing model in [**5**] says regarding the initial COVID-19 outbreak and subsequent lockdown in China. The software used in [**5**] does not seem to be available; this paper is accompanied by new public-domain software intended to implement the same model.

The new computations conclude that, with the minimum *R*_0_ allowed in the model of [**5**], China would have a 43-day period of hospitalizations being within 50% of peak, and a 53-day period of critical-care cases being within 50% of peak. For comparison, China’s reports of “severe cases” between 24 January and 28 March show only a 26-day period of “severe cases” being within 50% of peak.

This comparison is consistent with the theory that “intense” distancing cut China’s rate of new infections much more sharply than assumed in [**5**]. There was no way for [**5**] to rule out this theory, so [**5**] should have considered much smaller *R*_0_ values. This paper also covers various other problems with [**5**].

The theory stated in the previous paragraph should be treated with caution, for several reasons. The *hope* of strong effects from interventions creates a bias towards believing that such effects exist. Overly optimistic COVID-19 models have slowed down interventions (see, e.g., [**2**]), and newer models suggest that this slowness will turn out to have caused many unnecessary deaths; further policy decisions based on other unproven theories can be similarly disastrous. There are many other possible explanations for the relatively short period of “severe cases” in China: for example, perhaps increased contact tracing and wearing of masks helped reduce transmission; perhaps changes in treatment or in reporting policies reduced the number of reported “severe” cases.

Furthermore, the fragments of data considered here cannot reliably distinguish between, e.g., the theory that “intense” distancing reduces *R*_0_ by 90% and the theory that “intense” distancing reduces *R*_0_ by 99%. Policymakers trying to find the best combinations of non-pharmaceutical interventions to keep *R*_0_ low until a vaccine is available—for example, keeping it safely below 1, the “dance” explained in [**7**]—want to know much more regarding the impact of various types of distancing, the impact of encouraging widespread wearing of masks, etc. This paper should not be viewed as answering these questions; this paper merely disputes the overconfident answer given in [**2**].

The only policy recommendation in this paper is as follows. Governments should systematically allocate 5% of their daily COVID-19 testing capacity to testing people *chosen at random from census rolls*, with whatever incentives are necessary to create compliance (e.g., in the United States, a $100 reward to each person tested in this way). The resulting data regarding infection rates would help rule out incorrect theories regarding the prevalence of COVID-19, and would provide essential feedback regarding the impact of interventions. No claims of novelty are made regarding this recommendation.

## 2 The model

This section reviews [**5**]’s model of COVID-19 spread.

### 2.1. Introduction to SIR models

An “SIR model” works as follows. There are *N* people in the population. This population is partitioned into three sub-populations (“compartments”):

- Some people are “susceptible” people who have never been exposed to the disease. The fraction of people who are susceptible is called *S*, so in total there are *SN* susceptible people.
- Some people are “infectious” people who have been exposed to the disease and could be spreading it. The fraction of people who are infectious is called *I*, so in total there are *IN* infectious people.
- Some people are “recovered” people who have been exposed to the disease and are no longer spreading it. The fraction of people who are recovered is called *R*, so in total there are *RN* recovered people.

The fractions *S, I, R* have *S* + *I* + *R* = 1. These fractions change over time. Note that the literature often relabels *SN, IN, RN* as *S, I, R*, which also changes the equations shown below.

Each infectious person is assumed to take, on average, 1*/γ* days to recover, where *γ* is a parameter in the model. One can imagine several different ways to define the exact timing of recovery:

- Each infectious person recovers after exactly 1*/γ* days. (There is no variance in the recovery time.)
- Each infectious person has probability *γ* of recovering each day. (There is much more variance in the recovery time.)
- Each infectious person has probability *γ/*24 of recovering each hour.
- Et cetera.

An SIR model uses the limit of these possibilities, continuously compounding the probability of recovery. Mathematically, if there are no new infections, then this model says that the derivative *I*^*t*^(*t*) is *−γI*(*t*) where *t* is time measured in days, so *I*(*t*) = *I*(0) exp(*− γt*).

Meanwhile each infectious person is assumed to transmit the disease to an average of *β* people each day, spread uniformly at random through the entire population. This transmission affects only susceptible people: each infectious person infects, on average, *βS* people each day. The *IN* infectious people are assumed to infect separate people, a total of *βISN* people on average each day, increasing *I* at a rate *βIS*. This again is modeled as a continuous process:

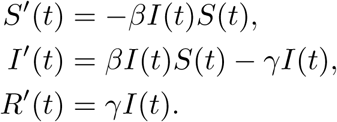

These ordinary differential equations are the definition of an SIR model. These equations are abbreviated “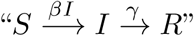”. Readers familiar with state-transition diagrams for discrete automata should observe that the notation here does not include self-loops, the default possibility of staying in the current compartment. Without doing the work of analyzing solutions to these equations, one can guess that each infectious person will transmit the disease to an average of *β/γ* people, and thus infect an average of (*β/γ*)*S* people, during the 1*/γ* days of an average infection. The ratio *β/γ* is called *R*_0_. This back-of-the-envelope calculation suggests that an epidemic with *R*_0_ *<* 1 will remain under control, while an epidemic with *R*_0_ *>* 1 will explode exponentially until *S* drops to 1*/R*_0_ (“herd immunity”).

One can see the same effects in the model: any nonzero rate of infectious people will increase (i.e., *I*^*t*^(*t*) *>* 0) as long as *βS*(*t*) *> γ*, i.e., *R*_0_*S*(*t*) *>* 1. Once *S*(*t*) drops to 1*/R*_0_, the quantity *I*(*t*) stops increasing. The remaining infectious people continue infecting more, pushing *S*(*t*) somewhat below 1*/R*_0_, but the recovery rate begins to reduce *I*(*t*).

### 2.2. General issues with SIR models

Real diseases seem to be less infectious at first but become more infectious after an incubation period; an SIR model does not account for this. An “SEIR model” tries to address this by adding an extra compartment for “exposed” people who are infected but not yet infectious:

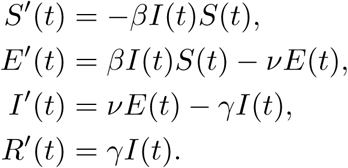

These equations are abbreviated “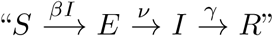”. The equations say that an exposed person takes, on average, 1*/ν* days to become infectious. Note that this delay slows down the progress of the epidemic, but also slows down the effect of interventions that are based only on observations of *I* and not on observations of *E*.

Real diseases also have different effects on different people—and, presumably, different levels of transmission—but SIR models and SEIR models do not account for this. The paper [**5**] tries to address this for COVID-19 by modeling three different levels of disease severity:

- Mild infections (“*I*_*R*_”) progress to recovery (“*R*_*R*_”).
- Moderate infections (“*I*_*H*_ “) progress to hospitalization (“*H*_*H*_ “), and then progress to recovery (“*R*_*H*_ “).
- Critical infections (“*I*_*C*_”) progress to hospitalization (“*H*_*C*_”), and then to critical care (“*C*_*C*_”), and then to recovery (“*R*_*C*_”).

The model thus has 11 compartments (including *S* and *E*). This makes the model sound more complicated than it actually is: one can compute the same results with just 7 compartments, namely *S, E, I* = *I*_*R*_ + *I*_*H*_ + *I*_*C*_, *H*_*H*_, *H*_*C*_, *C* = *C*_*C*_, and *R* = *R*_*R*_ + *R*_*H*_ + *R*_*C*_.

More broadly, when someone objects that an SIR/SEIR model is oversimplified, the standard response is to add more transitions to the model. For example:

- Perhaps observations show that immunity lapses. The standard response is to add an *R→ S* transition.
- Perhaps the observed variance of progression times for a disease is smaller than predicted in an SEIR model. The standard response is to add more stages of progression, such as *S →E*_1_*→ E*_2_ *→I*_1_ *→I*_2_ *→R*, narrowing the variance while preserving the average.
- Perhaps the period of infectiousness begins before the period of symptoms, and people with symptoms take more precautions to reduce transmission. The standard response—assuming, e.g., an *S → E*_1_ *→ E*_2_ *→ I*_1_ *→ I*_2_ *→ R* model—is to model a larger transmission speed from *I*_1_ than from *I*_2_. The terminology used for such a model distinguishes the “latent period”, meaning the non-infectious time spent in *E*, from the “incubation period”, meaning the pre-symptomatic time spent in *E* and *I*_1_.
- Part of the rationale for health-care workers to wear protective equipment is that hospitalized and critical-care patients—and the health-care workers themselves—are believed to be sources of infection. This is not accounted for in the model of [**5**]. The standard response would be to add further compartments to track infections of health-care workers.
- People are spread around the globe. Infections are sometimes carried by travelers or possibly by packages, but presumably infections are much more likely to spread locally. The standard response is to have local compartments, perhaps with different transmission rates that try to account for local factors such as population density and frequency of mask use.

All of these models depend heavily upon parameters such as *β* and *γ*. Small mistakes in estimating those parameters can easily produce vastly larger errors in predictions. This difficulty is exacerbated by the practice of adding more and more compartments to models, along with more and more dimensions in the parameter space, since the problem of computing parameters accurately from output data typically becomes exponentially more difficult as the dimension grows. Furthermore, modeling a discrete probabilistic process as a continuous deterministic process becomes increasingly difficult to justify as the number of compartments increases.

### 2.3. Parameters in [5]

In the model of [**5**], the speed (per day) of progression from being infectious (*I→ R* or *I→ H*_*H*_ or *I→ H*_*C*_, depending on the disease severity) is 1*/*5 (“*γ*”); the speed of progression from hospitalization to recovery (*H*_*H*_ *→ R*) for moderate cases is 1*/*8 (“*δ*_*H*_ “); the speed of progression from hospitalization to critical care (*H*_*C*_ *→ C*) is 1*/*6 (“*δ*_*C*_”); and the speed of progression from critical care to recovery (*C → R*) is 1*/*10 (“*ξ*_*C*_”). The speed of progression from being exposed to being infectious is not stated in [**5**] but appears to be modeled as 1*/*5.

In this model, a critical infection takes an average of 5+ 5+6 = 16 days before critical care, and an average of 16 + 10 = 26 days before the end of critical care. After a rapid burst of new infections, an intervention that drastically reduces new infections would not produce a peak of critical-care patients in this model until about three weeks later.

The model of [**5**] assumes that 95.6% of infections are mild while the other 4.4% require hospitalization. The procedures that have produced these numbers are not robust, as emphasized in [**3**], but this paper’s policy recommendation (see Section 1) would rapidly produce robust numbers. This is *not* an endorsement of the controversial idea from [**3**] of delaying serious interventions until robust numbers are available.

Within hospitalizations, the model of [**5**] assumes that 30% (i.e., 1.32% of total infections) require critical care.

There is one more parameter in the model of [**5**], the most important parameter: namely, *R*_0_, which in turn determines *β* = *γR*_0_. This is modeled in five stages:

- The wintertime *R*_0_ is modeled as either 2 or 2.5. Beware that the literature includes a wider range of estimates for *R*_0_, and underestimating *R*_0_ is dangerous.
- The summertime *R*_0_ is modeled as either 70% (more optimistic) or 100% (less optimistic) of the wintertime *R*_0_.
- The current-time *R*_0_ is modeled as a cosine curve from the wintertime *R*_0_ down to the summertime *R*_0_ and back up. The maximum is assumed to occur 3.8 weeks (26.6 days) before the end of the year.
- The distancing *R*_0_ takes 0%, 20%, 40%, or 60% away from the currenttime *R*_0_, reflecting four different hypotheses regarding the effectiveness of distancing.
- The final *R*_0_ is modeled as either the current-time *R*_0_ or the distancing *R*_0_, depending on whether distancing is “on” or “off”.

The paper analyzes the consequences in this model of a few different strategies for deciding when to turn distancing on or off.

To summarize, the differential equations for the model of [**5**] are as follows, where *R*_0_ is the final *R*_0_ defined above:

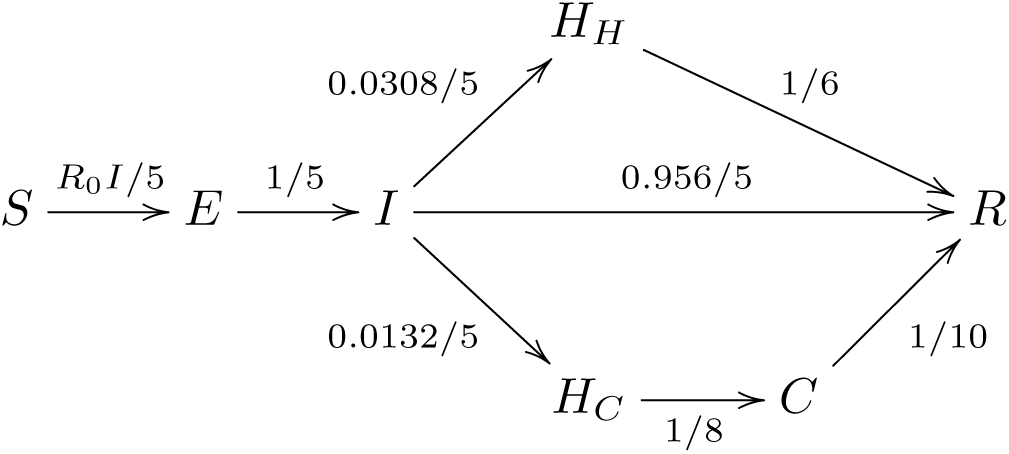

As mentioned earlier, this is presented with 11 compartments in [**5**], but these 7 compartments produce the same results in a simpler way.

### 2.4. Further issues with the model

According to Wikipedia, more than 10000 people in Italy have been reported dead with COVID-19 at the time of this writing, including more than 600 per day for each of the past 9 days. It is plausible (although not proven) that almost all of those deaths were not merely *with* COVID-19 but *caused by* COVID-19; and that if COVID-19 is not brought under control then it will cause millions of deaths worldwide within a year.

Perhaps many of these deaths can be avoided through, e.g., widespread mask usage, combined with several periods of distancing over the next year, followed by widespread vaccination. However, this cannot even be expressed, let alone analyzed, in the model of [**5**]:

- The model includes on/off distancing, but does not include masks or any other non-pharmaceutical interventions.
- The model does not include the possibility of future vaccination (*S →R*).
- The model does not include deaths. Dead people are treated as “recovered”— they have been exposed to the disease and are no longer spreading it. This simplification does not change the analysis of the spread of the epidemic, but the terminology is ethically questionable and presumably reduces the amount of attention given to one of the most important variables.

All of the parameters and distancing strategies considered in [**5**] result in mass COVID-19 infection within a few years—but a closer look shows that some scenarios have far fewer infections than others by mid-2021. In an extended model that accounts for deaths and the possibility of widespread vaccination, these scenarios would have far fewer deaths than other scenarios. There could be an even larger reduction in COVID-19 infections and deaths if non-pharmaceutical interventions are more effective than assumed in [**5**]. This again highlights the importance of understanding the actual impact of distancing, masks, etc.

## 3 What the model predicts for hypothetical future United States distancing

This section reviews, and disputes some of, (1) the calculations that [**5**] carries out within its model, and (2) the conclusions that [**5**] claims on the basis of these calculations.

### 3.1. Software engineering

https://cr.yp.to/2020/gigo-20200329.py, an attachment to this paper, is a Python script that produces (as PDF files) all of the graphs shown here.

As noted in Section 1, the software used in [**5**] does not seem to be available. It is easy to find software online for other SIR/SEIR/… models, including software that appears reasonably easy to adapt to the model of [**5**], but all of this software seems to depend on ODE-solving packages (e.g., `scipy.integrate.odeint`), and it is not clear how much review those packages have had from a safety-engineering perspective.

This paper’s Python script begins with a table of transitions, for example expressing 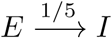 as follows:

~~~
(‘E’,’I’,1/5.0)
~~~

Transition speeds can be functions: for example, the transition

~~~
(‘S’,’E’,paperinitialpulse)
~~~

uses the following function

~~~
def paperinitialpulse(day,distancing):
 return 0.01/7 if day < startday+3.5 else 0
~~~

This appears to match the description in [**5**] of how the initial infection is modeled: “infection is introduced through a half-week pulse in the force of infection”. The height of this pulse is not stated in [**5**], but an earlier paper suggests 0.01 per week.

The most complicated transition is

~~~
(‘S’,’E’,paperbeta,’I’)
~~~

For 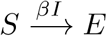. The function paperbeta is

~~~
def seasonalimprovement(day):
 summerimprovement = 0.3
 wintershift = 3.8*7 # peak of winter before end of December
 angle = 2*pi*(day+wintershift)/(7.0*seasonal)
 return summerimprovement*0.5*(1-cos(angle))
def paperbeta(day,distancing):
 R = R0
 if seasonal: R *= 1-seasonalimprovement(day)
 if distancing: R *= 1-0.6
 return R/5.0
~~~

where a careful reviewer needs to check every line against the model of *β* in [**5**]. The Python script then solves the corresponding differential equations from first principles: at each moment in time, use the table of transitions to compute the rate of change of each variable, and then compute

~~~
day += daydelta
for c in current:
 current[c] += change[c]*daydelta
~~~

to move to the next moment in time. The yes/no distancing variable is not differentiable and is instead updated by iterating the following function:

~~~
def paperstrategy(day,distancing):
if current[‘I’] >= 37.5/10000: distancing = 1
if current[‘I’] <= 10.0/10000: distancing = 0
return distancing
~~~

This matches the main distancing strategy highlighted in [**5**], where distancing is “turned ‘on’ when the prevalence of infection rose above … 37.5 cases per 10,000 people”, and off when the prevalence drops below 10.0 cases per 10,000 people. (“Infection” here appears to refer specifically to the *I* compartment, so it should have said “infectiousness”.)

If these calculations were performed in exact arithmetic (or interval arithmetic in higher and higher precision), rather than floating-point arithmetic, then the outputs over any compact interval would converge to the exact solution of the equations as `daydelta` converges down to 0. This does *not* imply that the outputs of the Python script are exact: the script uses floating-point arithmetic and a specific nonzero `daydelta`. More precisely, the script takes `daydelta` as 1/72 days (20 minutes), but decreases `daydelta` towards 0 when *I* approaches the 37.5/10000 or 10.0/10000 cutoffs. Some spot-checks suggest that smaller choices of `daydelta` produce visually identical graphs, but this is very far from a thorough error analysis.

Differential equations are normally solved by more complicated algorithms that are designed to obtain better tradeoffs between equation-solving time and accuracy. However, simpler algorithms are easier to review. The first 98 lines of this Python script include the transition tables, underlying functions, differential-equation solver, and recording the history of output data, using no libraries other than *π* and cos. The script then has more lines—which also need review!—for turning the data into graphs using `matplotlib`, a relatively complicated library.

### 3.2. Disputing the calculations in [5]

The paper [**5**] claims, within its model, that the (37.5, 10.0) distancing strategy explained above achieves the “goal of keeping the number of critical care patients below 0.89 per 10,000 adults” under the following assumptions: wintertime *R*_0_ = 2, and distancing achieves a 60% reduction in *R*_0_. This conclusion is stated for both the “seasonal and non-seasonal cases”, i.e., for summertime *R*_0_ being either 70% or 100% of the wintertime *R*_0_.

The conclusion is backed by a non-seasonal graph [**5**, Figure 3(A)] and by a seasonal graph [**5**, Figure 3(B)]. The seasonal graph comes close to exceeding 0.89 (in July 2021), but does not exceed it. The (37.5, 10.0) choice is close to, but not at, the edge of safe possibilities shown in [**5**, Figure S3]. The assumption that *R*_0_ = 2 is important: other graphs [**5**, Figures S6(A), S6(B)] say that this distancing strategy does not achieve the goal if *R*_0_ = 2.5.

**Fig. 3.3.**
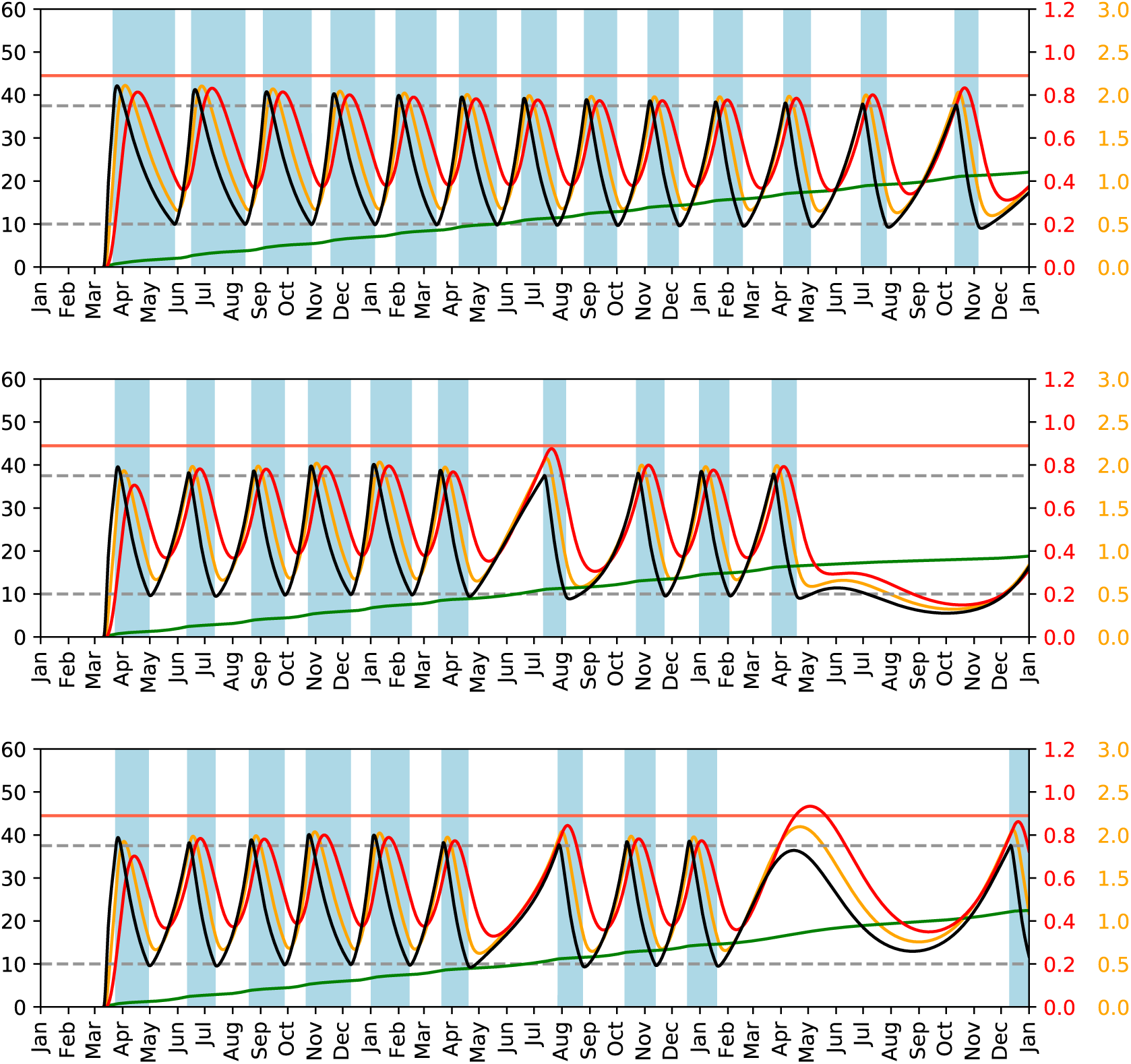
Model from [**5**], in the case *R*_0_ = 2, with 60% reduction from distancing. First plot: no seasonality. Second plot: model with a typo, 55-week 30% seasonality. Third plot: 52-week 30% seasonality. See text for further details.

**Fig. 3.4.**
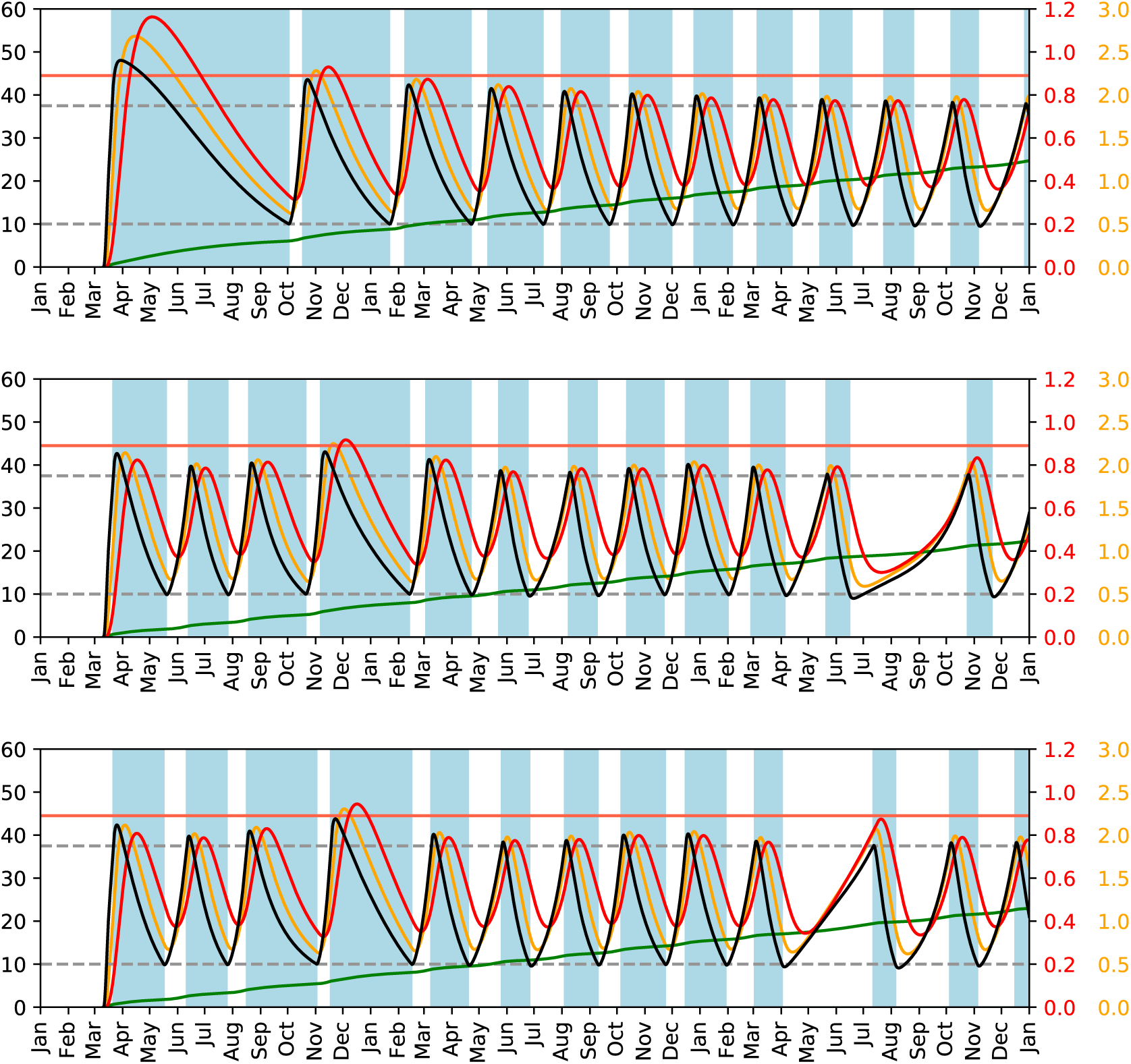
Model from [**5**], in the case *R*_0_ = 2.5, with 60% reduction from distancing. First plot: no seasonality. Second plot: model with a typo, 55-week 30% seasonality. Third plot: 52-week 30% seasonality. See text for further details.

**Fig. 3.8.**
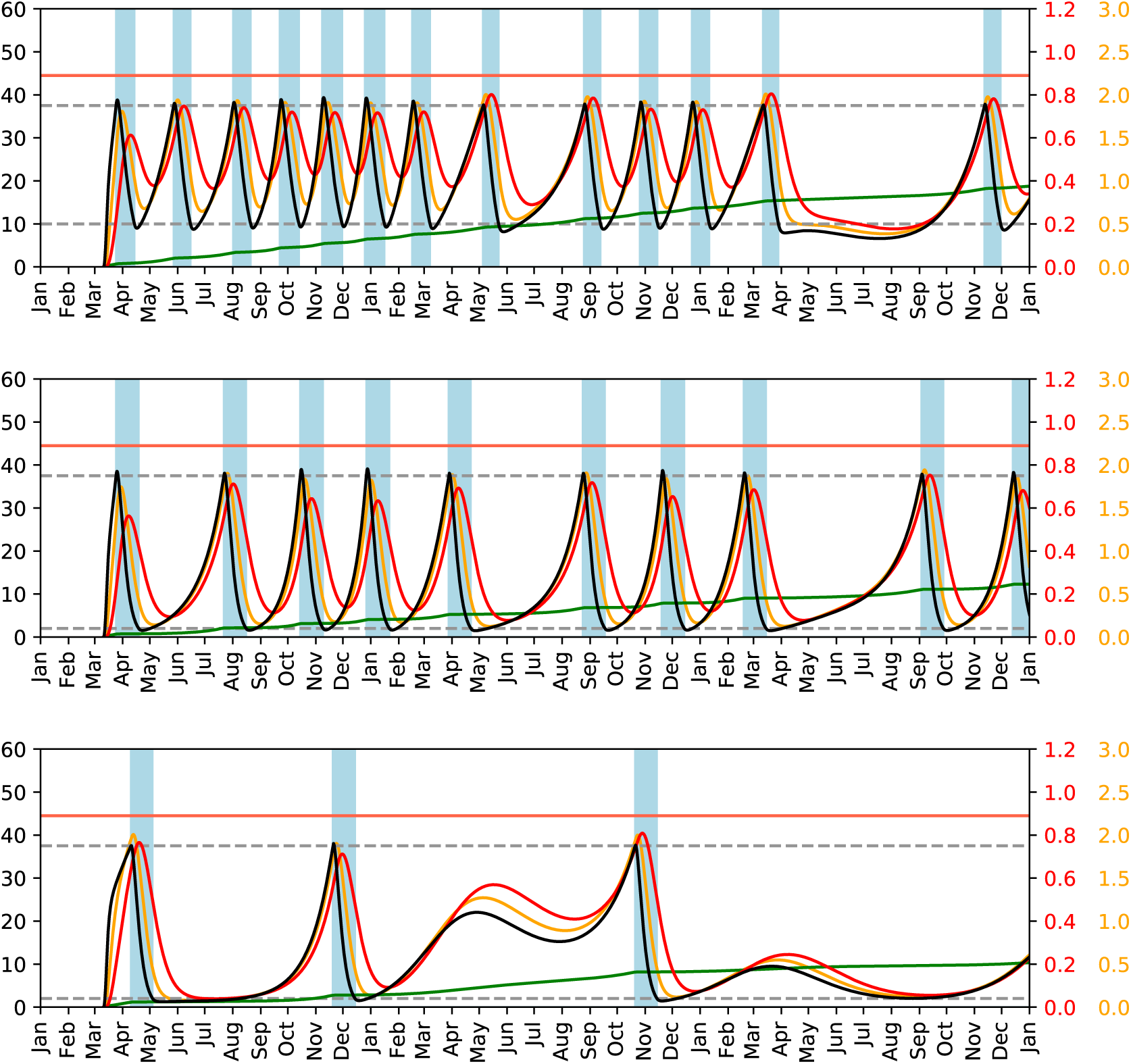
Extension of the model from [**5**] to consider the possibility of interventions further reducing *R*_0_. First plot: *R*_0_ = 2, 52-week 30% seasonality, 80% reduction from distancing, stopped at 10.0*/*10000. Second plot: *R*_0_ = 2, 52-week 30% seasonality, 99% reduction from distancing, stopped at 2.0*/*10000. Third plot: *R*_0_ = 1.5, 99% reduction from distancing, 52-week 30% seasonality, stopped at 2.0*/*10000. See text for further details.

Figure 3.3 shows three plots produced by this paper’s Python script with wintertime *R*_0_ = 2. The first plot is a recalculation of the non-seasonal [**5**, Figure 3(A)]. The black and red curves, as in [**5**, Figure 3(A)], show 10000*I* and 10000*C*; the top line shows a cutoff of 0.89 for 10000*C*, and the dotted lines show cutoffs of 37.5 and 10 for 10000*I*. The green curve shows 1 *−S* increasing to slightly above 0.4 by January 2023; this green curve matches [**5**, Figure 3(F)]. The orange curve shows 10000*H* = 10000*H*_*H*_ + 10000*H*_*C*_ (which is not plotted in [**5**]). As expected, each peak of infectiousness (black) triggers a later peak of hospitalizations (orange), followed by a peak of critical-care patients (red). These graphs go several months beyond the graphs in [**5**], so it is unsurprising that they show an extra period of distancing.

The second and third plots are two different recalculations of the seasonal [**5**, Figure 3(B)]. The second plot seems to match [**5**, Figure 3(B)]. The third plot looks the same at the beginning, but looks different in 2022, and in particular crosses the 0.89 line. The only difference between the models used in the second and third plots is as follows:

- One of the plots uses a 52-week seasonality period as specified in [**5**].
- The other plot is a calculation in a model that was modified through the introduction of a typographical error, replacing 52 with 55.

This illustrates how safety conclusions can be undermined by errors hidden inside epidemic-modeling software. This illustration is not hypothetical: the second plot, the one that seems to match [**5**, Figure 3(B)], is the one that includes the typographical error.

If the third plot here is correct then the (37.5, 10.0) safety claim in [**5**] is wrong. Occam’s razor suggests that the safety claim in [**5**], and the underlying calculations in [**5**], arise from the typographical error mentioned above.

In all of these models, the distancing trigger creates an immediate reduction in *E*, a less sharp reduction in *I* within a week, and a reduction in *C* in under a month. For comparison, peaks in *I* that occur without intervention (because 1*−S* has increased enough compared to *R*_0_) continue to trigger new infections for some time, and relatively large peaks in *C*. This is why the mid-2022 redversus-black gap in the third plot is relatively large compared to the other redversus-black gaps: a peak in *I* occurs just below the trigger for intervention. It should be clear that this variation can occur within this model, even under the assumptions that (1) there are mistakes in the third plot and (2) a corrected version of the same plot would not show this variation.

One can see this variation in gaps in [**5**, Figure S6(D)], but these phenomena— and their safety consequences—do not seem to have been noted in [**5**]. The error analyses in [**5**] were too limited to catch this error, and there is no evidence that the software was subjected to any double-checks or other review.

One can respond that 0.89 is not exceeded by *much* in the third plot, and that we should have enough critical-care capacity by then to handle this. Such a response is missing the broader point. If a change from 52 to 55 was not caught, why should one expect larger errors to be caught? Current practices in modeling epidemics obviously do not have adequate guarantees of the correctness of software claimed to be implementing models. The dangers of inaccuracies in computations are added to the dangers of inaccuracies in the models per se.

Email dated 25 Mar 2020 05:19:27 -0000 to the contact authors for [**5**] included a preliminary public version of this paper’s Python script, noted the discrepancy with [**5**, Figure 3(B)], noted the contradiction with the (37.5, 10.0) safety claim, and suggested posting software “for public review so that errors can be more easily located and corrected”. This email did not elicit a response. Considerable effort spent reverse-engineering [**5**, Figure 3(B)] eventually identified the theory of a 55 typographical error. The unavailability of the software continues to make the theory unnecessarily difficult to confirm.

As further checks on this paper’s Python script, Figure 3.4 includes the same calculations for *R*_0_ = 2.5. Compare the top two plots to [**5**, Figures S6(A), S6(B)].

**Fig. 4.2.**
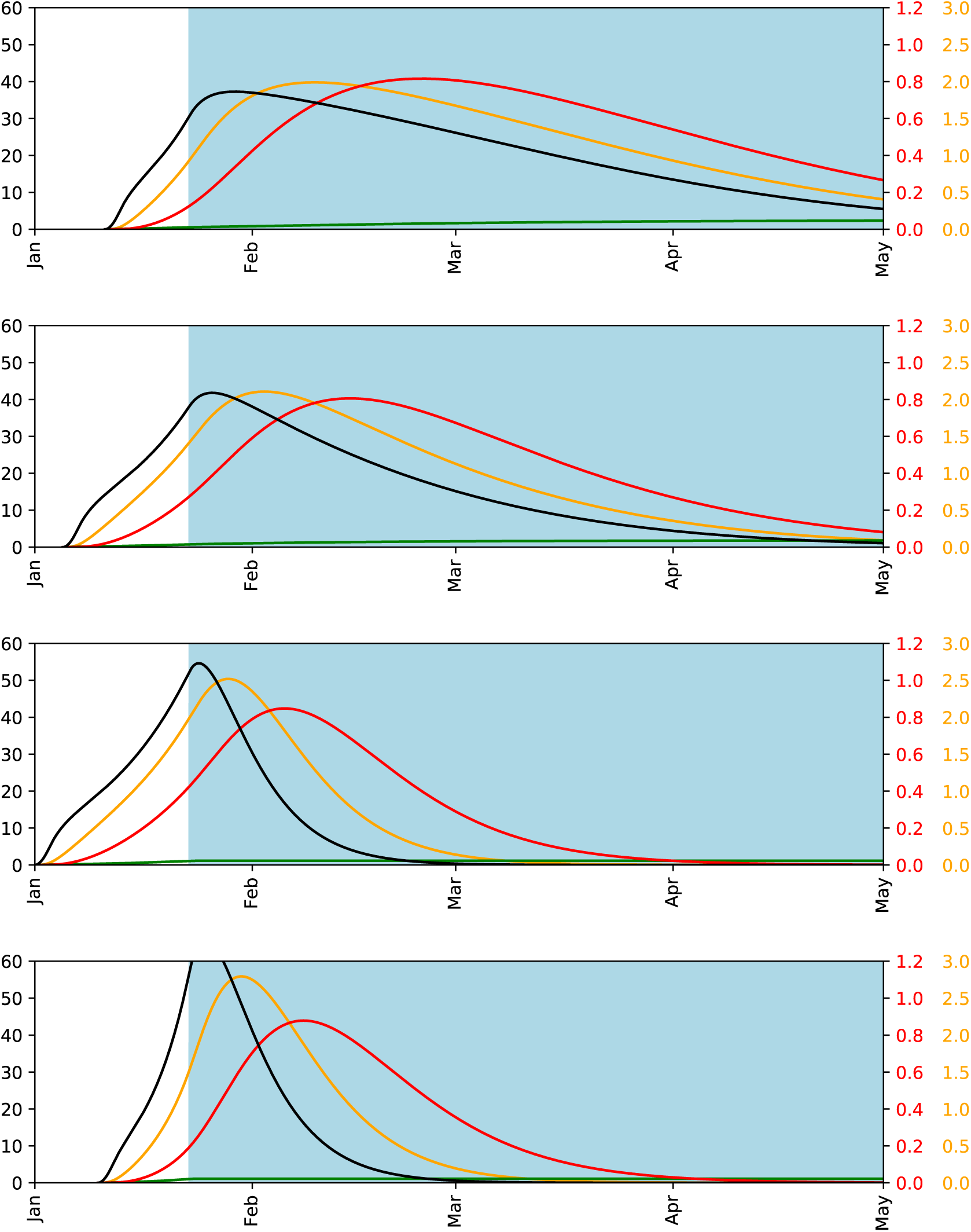
What the model from [**5**] predicts regarding a lockdown in China starting 23 January 2020. First plot: *R*_0_ = 2.5, with 60% reduction from distancing, with initial 2-day pulse starting 11 January 2020. Second plot: *R*_0_ = 2, with 60% reduction from distancing, with initial 2-day pulse starting 5 January 2020. Third plot: *R*_0_ = 2, in an extended model with 99% reduction from distancing, with initial 2-day pulse starting 1 January 2020. Fourth plot: *R*_0_ = 3.5, in an extended model with 99% reduction from distancing, with initial 2-day pulse starting 10 January 2020. See text for more details.

**Fig. 4.4.**
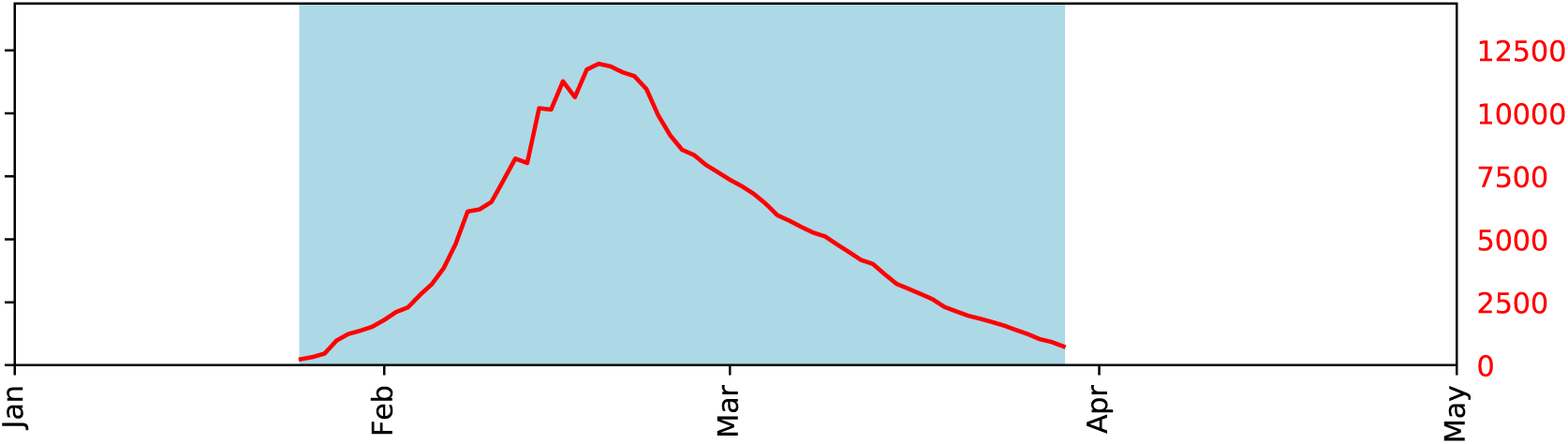
Red curve: Data from China’s NHC regarding “severe cases”. See text for further details.

### 3.5. Implementability of distancing strategies

Recall that this distancing strategy is triggered by *I* crossing particular cutoffs, namely 37.5*/*10000 and 10.0*/*10000. A policymaker considering this strategy immediately runs into the problem that the real-world *I* is unknown at each moment. (This is separate from the question of whether the strategy is safe; see above.)

The paper [**5**] claims that, to “implement an effective intermittent social distancing strategy”, it will be “necessary to carry out widespread surveillance to monitor when the prevalence thresholds that trigger the beginning or end of distancing have been crossed”.

Collecting data regarding COVID-19 prevalence is valuable for other reasons, and is the only policy recommendation in this paper. This does not imply, however, that such data collection would enable implementation of the strategy in [**5**] with a useful level of accuracy. An error analysis is required here—accounting for lags in testing, errors in testing, possible correlations between infectiousness and non-compliance, etc.—but does not appear in [**5**].

Furthermore, no justification is provided in [**5**] for the claim that this data collection is “necessary” for a distancing strategy to be “effective”. Imagine policymakers today implementing a simple one-month-on-one-month-off distancing strategy to be adjusted later; this is effective in some of the scenarios covered by the paper’s model, contradicting the blanket claim of ineffectivity. Perhaps this simple strategy is unimplementable for cost reasons or political reasons, but this does not justify the claim that the strategy is ineffective.

### 3.6. The effect of increased critical-care capacity

The obvious way to handle the possibility of overruns in critical-care capacity is to increase the criticalcare capacity. This is not controversial.

What is much more controversial is the idea of actively *trying to exploit all available critical-care capacity*, tuning the amount of distancing to almost—but not quite—overload this capacity. One reason this is controversial is that such tuning is prone to error, whether the errors come from miscalculations within models (as illustrated above) or from the models being wrong; see, e.g., [**8**]. Another, more fundamental, reason this is controversial is that less distancing means more infections. The claim that these infections are inevitable *might* be correct but (1) is not justifiable from the available data and (2) does not justify policy decisions that make the infections happen *now*.

The paper [**5**] claims that increasing critical-care capacity “allows population immunity to be accumulated more rapidly, reducing the overall duration of the epidemic and the total length of social distancing measures”. It is not ethical to say that exploiting increased critical-care capacity in this way is “allowed” without mentioning that this has the side effect of more infections now—including painful hospitalizations and deaths that would have been avoided by more distancing. Furthermore, the predictions made by the model of [**5**], in particular regarding the epidemic duration and the amount of distancing, are not robust against modifications to the model that account for more effective interventions and the possibility of widespread mid-2021 vaccinations.

The paper [**5**] also claims, in its “Summary” at the top, that “Intermittent efforts require greater hospital capacity”. This is contradicted by, e.g., [**5**, Figure 3(B)], which claims—for *R*_0_ = 2 with seasonality—that the capacity of 0.89 critical-care beds per 10000 adults would not be overrun with (roughly) halftime intermittent distancing in 2020 and less distancing in subsequent years. The second claim appears to be based on a miscalculation, as noted above, but correcting this miscalculation and adding more distancing in subsequent years would again contradict the “require” claim.

This criticism of various unjustified claims in [**5**] should not be interpreted as opposition to the idea of increasing hospital capacity. Current news reports and simple extrapolations are consistent with the theory that the United States will already need more than 0.89 critical-care beds per 10000 adults in April, as a direct result of inadequate initial interventions in March.

### 3.7. The possibility of interventions being more effective than assumed in the model

The first sentence of the summary of [**5**] is as follows: “One-time distancing results in a fall COVID-19 peak.” As for intermittent distancing, the paper’s lead author [**4**] summarizes the paper’s main quantitative conclusion as follows:

> Intermittent social distancing can prevent critical care capacity from being exceeded but such measures may be required for 12-18 months. Depending on R0 and the amount of seasonality, social distancing must be ‘on’ for as little as 25% but up to 70% of the time.

The paper itself says that staying below 0.89 requires “social distancing measures to be in place between 25% … and 70% of that time” (into 2022). These claims *could* be correct, but they go beyond what is shown in the paper.

Concretely, the paper shows *within its model* that a single month of distancing now will not prevent mass infection by October; and that extending this to, say, three months will not prevent mass infection by the end of the year. The paper also shows *within its model* that a particular intermittent-distancing strategy, applied between 25% and 70% of the time, comes close to 0.89/10000 criticalcare patients again and again. It is reasonable to conjecture that, within this model, every intermittent-distancing strategy with fewer than 6 months of total distancing will break the 1/10000 barrier.

However, all of these conclusions depend upon the exact values of *R*_0_ during and after distancing. Perhaps the effect of distancing is stronger than assumed in [**5**]. Perhaps other interventions—increased hand-washing, increased wearing of masks and mask substitutes, etc.; see [**7**] for many more possibilities—will keep *R*_0_ below 1 after one-time distancing.

As examples of how heavily the results depend upon hypotheses regarding the effectiveness of interventions, Figure 3.8 shows what happens with the following strategies and extensions of the model: (1) distancing reduces *R*_0_ by 80%; (2) distancing is turned off at 2.0*/*10000, and reduces *R*_0_ by 99%; (3) same but with *R*_0_ starting at 1.5 instead of 2, as a model of non-distancing interventions reducing transmission by 25%. The clear differences between the graphs again highlight how important it is to understand the actual impact of interventions.

## 4 What the model predicts for actual China distancing

This section returns to the claim in [**5**] that a 60% reduction in *R*_0_ is “on par with the reduction in *R*_0_ achieved in China through intense social distancing measures”. As noted in Section 1, this claim is incorrectly attributed to [**1**], and is not otherwise justified in [**5**]. Sections 2 and 3 have highlighted the importance of understanding what *R*_0_ can actually be achieved.

### 4.1. Computations in the model

Figure 4.2 contains four more plots produced by the same Python script used in Section 3. Each plot stretches over a 4-month period, includes 52-week seasonal forcing, and includes “intense” distancing starting on 23 January 2020.

The second plot takes the wintertime *R*_0_ to be 2.0, the most optimistic possibility allowed in [**5**]. “Intense” distancing is assumed to reduce *R*_0_ by 60%, again the most optimistic possibility allowed in [**5**].

The first (less optimistic) plot takes the wintertime *R*_0_ to be 2.5. The third (more optimistic) plot takes the wintertime *R*_0_ to be 2.0 and uses an extended model where “intense” distancing has more of an effect, reducing *R*_0_ by 99%. The fourth plot is like the third but takes the wintertime *R*_0_ to be 3.5.

In each plot, there is an initial 2-day pulse of exposure. This is chosen shorter than the half-week pulse from [**5**] to limit the impact of the pulse time upon the width of the resulting peaks. The pulse starts on 11 January, 5 January, 1 January, and 10 January respectively; these dates are chosen so that the red curves have approximately the same peak heights, simplifying comparison of other features of the curves.

### 4.3. Reported “severe cases” from China

https://cr.yp.to/2020/nhc-20200329.py is a Python script that is intended to plot, for each day, the number of cases reported by China’s NHC as being “severe” on that day. The data incorporated into the script was extracted manually, with considerable help from Google Translate, from http://www.nhc.gov.cn/yjb/pzhgli/new_list.shtml, the primary source of China COVID-19 case counts cited by Wikipedia. Double-checking the data, and other aspects of the script, would be useful. The output of the script is shown in Figure 4.4.

The reason for selecting reports of “severe” cases, rather than (e.g.) reports of “confirmed” cases, is the common-sense guess that more severe cases are more likely to be tested and reported, hopefully producing a curve close to reality. However, there could still be biases in the testing or reporting procedures, and the NHC reports do not state the exact definition of a “severe” case.

### 4.5. Comparing the model’s predictions to the reports

None of the red curves in Figure 4.2 is a convincing fit to the reports in Figure 4.4. Specifically, each of the curves in Figure 4.2 deviates from Figure 4.4 in at least one of the following two metrics:

- Width of the peak: e.g., number of days during which the red curve is at least half of its maximum. This width is 73.7, 53.4, 32.2, 30.2 in Figure 4.2, and just 26 in Figure 4.4.
- Position of the maximum. This position is at day 55.1, 44.9, 35.6, 38.3 (counting from 0 at the beginning of the year) in Figure 4.2, and at day 49 in Figure 4.4.

This raises the question of how to build a model that is not obviously inconsistent with the NHC reports—while at the same time avoiding the clear danger of overfitting.

There are several contributing factors to the widths of the peaks in Figure 4.2. In the first plot, wintertime *R*_0_ = 2.5 is reduced by distancing to 1, so the epidemic is controlled primarily by seasonal forcing (plus a gradual increase in 1*− S*, the green curve). This is not a large effect, so new infections continue to occur for a long time. This phenomenon is smaller in the second plot, with wintertime *R*_0_ = 2, and much smaller in the third and fourth plots.

The width of the *E* peak (not plotted) then produces a somewhat wider *I* peak (the black curve), since *E → I* has a high variance. This in turn produces a somewhat wider *H* peak (the orange curve) followed by a somewhat wider *C* peak (the red curve).

As noted in Section 2, SEIR models are easily adjusted to match observations of lower variance in an individual’s disease progression. For example, the analysis of [**6**] indicates that the COVID-19 incubation time has standard deviation only about half of its mean. Modifying the model of [**5**] to include more *E* stages (while preserving the mean) would reduce the width of the resulting *I, H*, and *C* peaks, producing a sharper increase towards the maximum and the a sharper decrease from the maximum. It is also easy to adjust the model of [**5**] to delay the *C* peak.

However, a large part of the width of the peak in the first and second plots in Figure 4.2 arises directly from the assumption that *R*_0_ is large even after distancing. It is not obvious how to reconcile the model of [**5**] with the reports from Figure 4.4 without dropping this assumption.

This analysis should not be interpreted as confidently concluding that [**5**] is wrong in claiming that China achieved only a 60% reduction in *R*_0_. One can imagine various ways that the claim could still be correct. However, [**5**] is not justified in the level of confidence that it states in this claim, and is not justified in the level of confidence that it states in the conclusions that rely on this claim.

## Data Availability

The URLs are for open-source Python scripts that reproduce all computations and graphs in this paper. The second script includes the relevant NHC data.

https://cr.yp.to/2020/gigo-20200329.py

https://cr.yp.to/2020/nhc-20200329.py

## A Change log

Chronological order.

**2020**.**03**.**29:** First public version.

Corrected typo in parenthetical note regarding definition of “*δ*_*C*_”: the note said “*H*_*H*_ *→C*” instead of the correct “*H*_*C*_*→C*”. (This typo did not appear in this paper’s differential equations, and did not appear in this paper’s software, but illustrates the importance of double-checking everything.)

Broadened “sensitivity analysis” to “error analysis”, for readers who expect “sensitivity” to refer specifically to the sensitivity of a test. Also changed “are sensitive to” to “depend upon”.

In the list of illustrations of SIR/SEIR oversimplifications, added note on one way to model an incubation period longer than a latent period, and expanded description of local models to include simpler models that do not vary in local transmission rates.

Added *S* and *R* explicitly around *E*_1_*→ E*_2_*→ I*_1_*→ I*_2_ for clarity.

Replaced the wording “accurate fitting” with the wording “the problem of computing parameters accurately from output data”, to make clear that the accuracy metric here is the accuracy of the resulting parameters.

Added comment that the notation here does not include self-loops, for readers familiar with state-transition diagrams for discrete automata.

Improved spacing for code excerpts.

**2020**.**03**.**30:** This version.

## References

[1] Roy M. Anderson, Hans Heesterbeek, Don Klinkenberg T Déirdre Hollingsworth, How will country-based mitigation measures influence the course of the COVID-19 epidemic?, The Lancet 395:10228 (2020), 931–934. URL: https://www.thelancet.com/journals/lancet/article/PIIS0140-67362030567-5/fulltext.Citations in this document: §1, §1, §1, §4.

[2] Martin Enserink, Kai Kupferschmidt, Mathematics of life and death: How disease models shape national shutdowns and other pandemic policies, Science Magazine (2020). URL: https://www.sciencemag.org/news/2020/03/mathematics-life-and-death-how-disease-models-shape-national-shutdowns-and-other. Citations in this document: §1,§1

[3] John P. A. Ioannidis, A fiasco in the making? As the coronavirus pandemic takes hold, we are making decisions without reliable data (2020). URL: https://www.statnews.com/2020/03/17/a-fiasco-in-the-making-as-the-coronavirus-pandemic-takes-hold-we-are-making-decisions-without-reliable-data/.Citations in this document: §2.3, §2.3.

[4] Stephen Kissler, Intermittent social distancing can prevent critical care capacity from being exceeded but such measures may be required for 12-18 months. Depending on R0 and the amount of seasonality, social distancing must be ‘on’ for as little as 25% but up to 70% of the time, tweet (2020). URL: https://twitter.com/StephenKissler/status/1242106525902090241. Citations in this document: §3.7.

[5] Stephen Kissler, Christine Tedijanto, Marc Lipsitch, Yonatan Grad, Social distancing strategies for curbing the COVID-19 epidemic (2020). URL: https://dash.harvard.edu/handle/1/42638988. Citations in this document: §1, §1, §1, §1, §1, §1, §1, §1, §1, §1, §1, §1, §2, §2.2, §2.2, §2.3, §2.3, §2.3, §2.3, §2.3, §2.3, §2.3, §2.3, §2.4, §2.4, §2.4, §3, §3, §3.1, §3.1, §3.1, §3.1, §3.1, §3.1, §3.2, §3.2, §3.2, §3.2, §3.2, §3.2, §3.3, §3.3, §3.2, §3.2, §3.2, §3.2, §3.2, §3.2, §3.2, §3.2, §3.2, §3.2,§3.2, §3.2, §3.2, §3.2, §3.2, §3.2, §3.2, §3.2, §3.2, §3.4, §3.4, §3.5, §3.5, §3.5, §3.5,§3.6, §3.6, §3.6, §3.6, §3.6, §3.7, §3.8, §3.8, §3.7, §4, §4, §4.2, §4.2, §4.1, §4.1, §4.1,§4.5, §4.5, §4.5, §4.5, §4.5.

[6] Ke Men, Xia Wang, Li Yihao, Guangwei Zhang, Jingjing Hu, Yanyan Gao, Henry Han, Estimate the incubation period of coronavirus 2019 (COVID-19) (2020). URL: https://www.medrxiv.org/content/10.1101/2020.02.24.20027474v1. Citations in this document: §4.5.

[7] Tomas Pueyo, Coronavirus: the hammer and the dance (2020). URL: https://medium.com/@tomaspueyo/coronavirus-the-hammer-and-the-dance-be9337092b56. Citations in this document: §1, §3.7.

[8] Nassim Nicholas Taleb, Lesson from a long experience with model blowups:@neil ferguson, if you need a model w/”thousands of lines”, this is not a model useable for real world risk & decisions–rather something with the FRAGILITY of a house built with matches to impress some tenure committee, tweet (2020). URL: https://twitter.com/nntaleb/status/1242235586578190338. Citations in this document: §3.6.

